# Recommendations for COVID-19 Vaccination in People with Rheumatic Disease: *Developed by the Singapore Chapter of Rheumatologists*

**DOI:** 10.1101/2021.03.01.21252653

**Authors:** Amelia Santosa, Chuanhui Xu, Thaschawee Arkachaisri, Kok Ooi Kong, Aisha Lateef, Tau Hong Lee, Keng Hong Leong, Andrea Hsiu Ling Low, Melonie K Sriranganathan, Teck Choon Tan, Gim Gee Teng, Bernard Yu-hor Thong, Warren Fong, Manjari Lahiri

**Author notes:** Correspondence: Dr Manjari Lahiri, Division of Rheumatology, Department of Medicine, National University Hospital, Singapore, 1E Kent Ridge Road, Singapore 119228.

## Abstract

**Aim:** People with rheumatic diseases (PRD) remain vulnerable in the era of the COVID-19 pandemic. We formulated recommendations to meet the urgent need for a consensus for vaccination against SARS-CoV-2 in PRD.

**Methods:** Systematic literature reviews were performed to evaluate (1) outcomes in PRD with COVID-19; (2) efficacy, immunogenicity and safety of COVID-19 vaccination; and (3) published guidelines/recommendations for non-live, non-COVID-19 vaccinations in PRD. Recommendations were formulated based on the evidence and expert opinion according to the Grading of Recommendations Assessment, Development and Evaluation methodology.

**Results:** The consensus comprises two overarching principles and seven recommendations. Vaccination against SARS-CoV-2 in PRD should be aligned with prevailing national policy and should be individualized through shared decision between the healthcare provider and patient. We strongly recommended that eligible PRD and household contacts be vaccinated against SARS-CoV-2. We conditionally recommended that the COVID-19 vaccine be administered during quiescent disease if possible. Immunomodulatory drugs, other than rituximab, can be continued alongside vaccination. We conditionally recommended that the COVID-19 vaccine be administered prior to commencing rituximab if possible. For patients on rituximab, the vaccine should be administered a minimum of 6 months after the last dose and/or 4 weeks prior to the next dose of rituximab. Post-vaccination antibody titres against SARS-CoV-2 need not be measured. Any of the approved COVID-19 vaccines may be used, with no particular preference.

**Conclusion:** These recommendations provide guidance for COVID-19 vaccination in PRD. Most recommendations in this consensus are conditional, reflecting a lack of evidence or low-level evidence. (words 247)

## Introduction

The global novel coronavirus disease 2019 (COVID-19) pandemic has posed many uncertainties among physicians treating people with rheumatic diseases (PRD). Such patients are considered high risk due to their diseases and the immunosuppressive nature of their medications. A recent meta-analysis demonstrated that PRD had a two-fold risk of COVID-19 compared to control patients.^1^ In addition, PRD with COVID-19 had a higher fatality rate and were at significant risk of suffering poor outcomes such as the need for hospitalization, care in the intensive care unit (ICU) and mechanical ventilation^2,3^

Various candidate vaccines against severe acute respiratory syndrome coronavirus-2 (SARS-CoV-2) are in development. The first three COVID-19 vaccines to receive Emergency Use Authorisation (EUA) from the United States Food and Drug Administration (US FDA), the Pfizer-BioNTech® COVID-19 vaccine (BNT162b2), the Moderna® COVID-19 vaccine (mRNA-1273) and the Johnson & Johnson^®^ vaccine (JNJ-78436735), reported good vaccine efficacy at 95%, 94.1% and 66.9%, respectively.^4-6^ However, patients on immunosuppressive therapy were excluded from all three trials. Additionally, patients with autoimmune diseases were excluded from two of the trials, and only 62 PRD (0.3% of the total study population, but without detailed information) were included in the treatment arm of the Pfizer-BioNTech® trial. Thus, there is a paucity of evidence for PRD and their managing physicians to guide COVID-19 vaccination in this population.

The Singapore Health Sciences Authority (HSA) has approved the Pfizer-BioNTech^®^ and Moderna^®^ COVID-19 mRNA vaccines via the Pandemic Special Access Route, and the Ministry of Health, Singapore (MOH) Expert Committee on COVID-19 Vaccination (EC19V) has published recommendations for their use ^7,8^ with other vaccines to be evaluated at a later date. Worldwide, four additional vaccines, namely from Gamaleya Research Institute of Epidemiology and Microbiology (Gam-COVID-Vac or Sputnik V^®^) ^9^, Oxford-Astra-Zeneca^®^ (AZD1222) ^10^, Novartis (Novavax^®^ or NVX-CoV2373) ^11^ and Bharat Biotech (BB-152 or Covaxin^®^) ^12^, have so far published or announced interim Phase 3 efficacy data and are either already authorised or expected to apply for EUA in several countries. In this consensus recommendation, the Chapter of Rheumatologists, College of Physicians, Academy of Medicine, Singapore seeks to address questions regarding the suitability of COVID-19 vaccination in PRD and provide consensus recommendations on COVID-19 vaccination among PRD.

## Methods

A Core Working Group (CWG) was established (AS, CX, WF, ML). Members of the CWG reviewed published primary clinical trials and performed a systematic literature review to answer four research questions. Where appropriate, in lieu of a systematic review of the primary literature, international best practice guidelines and recommendations from rheumatology societies on vaccinations in PRD were reviewed. Other academic bodies’ recommendations for COVID-19 vaccination and other non-live, non-COVID-19 vaccinations in PRD and / or immunocompromising conditions were also considered. The CWG developed draft recommendations for rating by an invited task force panel (TFP). A modified Delphi approach, similar to what has been applied by other organizations, was used^13,14^ The TFP (TA, KOK, AL, THL, KHL, AHLL, MKS, TCT, GGT, BYT) comprised eight locally recognized adult rheumatologists from public and private healthcare institutions in Singapore, one paediatric rheumatologist and one infectious diseases specialist. A conflict-of-interest declaration was required from all members of the CWG and TFP prior to the consensus process. All members declared no conflict of interest.

### Review of the literature

The CWG sent out preselected topics to the TFP and sought their input on additional clinically important topics. Considering the TFP’s input, the CWG selected the following core topics relevant to clinical decision-making for COVID-19 vaccination:

#### 1. Are PRD at increased risk of adverse outcomes from COVID-19?

A recent systematic review and meta-analysis of global data showed that PRD remain vulnerable, with substantial rates of severe outcomes.^3^ The overall rates of hospitalization, oxygen support, ICU admission and fatality among COVID-19 infected patients with rheumatic diseases were 58% (95% confidence interval (CI) 48% - 67%), 33% (95% CI 21% - 47%), 9% (95% CI 5% - 15%) and 7% (95% CI 3% - 11%), respectively, which are comparable with data from the COVID-19 Global Rheumatology Alliance (GRA) physician registry. The fatality rate was higher both in this meta-analysis and the COVID-19 GRA (7.0% and 6.7%, respectively) than that (3.4%) of general population infected with COVID-19 in the WHO database, although age, gender and comorbidities were not matched.^3^ D’Silva *et al* reported a higher risk of hospitalization, ICU admission, mechanical ventilation, acute kidney injury, renal replacement therapy and death based on TriNetX, a multi-center research network with real-time electronic health record data across 35 healthcare organizations in the US.^15^ The authors concluded that these outcomes were likely mediated by a higher comorbidities burden in PRD, such as hypertension, diabetes mellitus, chronic kidney disease and asthma.

#### 2. Are existing approved vaccines against SARS CoV2 safe, immunogenic and efficacious in PRD?

Two mRNA vaccines are currently approved by the US FDA and Singapore HSA. It is known that selected DNA and RNA molecules have the unique property to activate the immune system, through activation of Toll-like receptors.^16^ It has been shown that the innate immune response would be suppressed by nucleoside modification of RNA, as the innate immune system detects RNA lacking nucleoside modification as a means of selectively responding to bacteria or viruses.^17,18^ Both mRNA COVID-19 vaccines from Pfizer/BioNTech^®^ and Moderna^®^ are nucleoside-modified RNA. Thus, the risk of autoimmune disease flare after receiving mRNA COVID-19 vaccine may more likely result from the adaptive immune response to spike protein synthesized by mRNA, rather than the innate immune response to nucleoside-modified RNA. Theoretically, this is no different from the risk from other protein / conjugate vaccines, which have been in use for many years and have been confirmed to be safe in PRD.

There were 62 (0.3%) participants who had rheumatic disease and received BNT162b2 mRNA COVID-19 vaccine in the Pfizer/BioNTech^®^ trial.^4^ No flare of autoimmune disease was reported. Certainly, larger sample size and long-term follow-up studies are needed to further ascertain the risk of flares in autoimmune diseases.

Other vaccine strategies, including inactivated virus vaccines (such as the CoronaVac developed by Sinovac^® 19^ and Covaxin^®^ developed by Bharat Biotech ^12^), virus vector vaccines (such as the COVID-19 vaccines by AstraZeneca^® 10^, the Johnson and Johnson vaccine ^6^ and the Sputnik V^®^ Russian vaccine by Gamaleya ^9^) and protein subunit vaccines (such as the Novavax^®^ vaccine ^11^) similarly provide little data in PRD. Pertinent information from primary COVID-19 vaccine trials to date are summarized in Table 1.^4-6,9,10,12,19,20^

**Table 1:** Primary COVID-19 vaccine trials

|  | Pfizer-BioNTech®<sup>4</sup> | Moderna®<sup>5</sup> | Sinovac®<sup>19</sup> | Oxford-Astra<br>Zeneca®<sup>10</sup> | Gamaleya®<sup>9</sup> | Johnson & Johnson®<br>6 | Novartis<sup>20</sup> | Bharat Biotech<sup>12</sup> |
| --- | --- | --- | --- | --- | --- | --- | --- | --- |
| Trial Sites | US, Brazil, Argentina, South Africa, Germany, Turkey | United States | China | UK, Brazil, South Africa | Russia | US, Central & South America, South Africa | UK, South Africa | India |
| MOA | Lipid nanoparticle–formulated, nucleoside-modified mRNA |  | Adsorbed SARS-CoV-2 (inactivated) vaccine | Replication deficient viral vector with SARS COV2 spike protein |  |  | Adjuvant recombinant protein particle | Whole-virion inactivated SARS-CoV-2 vaccine with a toll-like receptor 7/8 agonist molecule adsorbed to alum |
| Storage | Freezer -70°C | Freezer -15 to -25°C | Refrigerator 2 to 8°C | Refrigerator 2 to 8°C | Freezer -18°C | Refrigerator 2 to 8°C | Refrigerator 2 to 8°C | Refrigerator 2 to 8°C |
| Dosing | Two 30ug (0.3ml) IM doses 21 days apart | Two 100ug (0.5ml) IM doses 28 days apart | Two doses 2 weeks apart | Two doses 4-12 weeks apart | Two (0.5ml) IM doses 21 days apart | Single dose | Two (0.5ml) IM doses 21 days apart | Two 6ug IM doses 28 days apart |
| Inclusion | Adults (>16yrs) | Adults (≥18yrs) | Adults (≥18yrs) | Adults (≥18yrs) | Adults (≥18yrs) | Adults (≥18yrs) | Adults (18-84yrs) | Adults (18-98yrs) |
| Relevant Exclusions | Immunodeficient state and use of immunosuppressant medication. | Autoimmune disease Immunodeficient state and use of immunosuppressant medication within the past 6 months. | Autoimmune disease Immunodeficient state and use of immunosuppressant medication within the preceding 3 months | Autoimmune disease Immunodeficient state and use of immunosuppressant medication within the past 6 months. | Immunodeficient state and use of immunosuppressant medication within the past 3 months. | Immunodeficient state and use of immunosuppressant medication. | Autoimmune disease Immunodeficient state and use of immunosuppressant medication within the preceding 3 months | NA |
| N* | 37,706 | 28,207 | 13,060 | 11,636 | 19,866 | 43,783 | 15,000 | 25,800 |
| Follow up (median) after last dose | 2 months | 64 days | NA | 2 months | 27 days | 8 weeks | NA | NA |
| Asian participants | 1608 (4.3%) | 1382 (4.6%) | NA | 517 (4.4%) | 286 (1.4%) | 3.5% | NA | 100% |
| Comorbidities | 20.5% | 22.3% | NA | 24.7% | 24.8% | 41% (including obesity) | NA | 17.4% |
| PRD | 62 (0.3%) | Excluded | NA | Excluded | Likely excluded | Allowed, likely none included | NA | NA |
| Elderly | >55yrs (42.2%) | >65yrs (25.3%) | NA | >55yrs (12.2%) | >60yrs (10.8%) | >65yrs (20.4%) | >65yrs (27%) | >60 yrs (9.4%) |
| Outcomes | 8 vs 162 cases of symptomatic and PCR confirmed COVID19, vaccine efficacy 95% | 11 vs 185 cases of symptomatic and PCR confirmed COVID19, vaccine efficacy 94.1% | NA | 30 vs. 101 cases of symptomatic and PCR confirmed COVID19, vaccine efficacy 70.4% | 16/14964 vs. 62/4902 cases of symptomatic and PCR confirmed COVID19, vaccine efficacy 91.6% | 116/19514 vs 348/19544 moderate / severe PCR confirmed COVID19, | 6 vs. 56 cases of symptomatic and PCR confirmed COVID19, vaccine efficacy 89.3% | 7 vs. 36 cases of symptomatic and PCR confirmed COVID19, vaccine efficacy 80.6% |

|  |  |  |  |  |  | vaccine efficacy<br>66.9% |  |  |
| --- | --- | --- | --- | --- | --- | --- | --- | --- |
| Common adverse events | Expected reactogenicity: fatigue, headache. Rare anaphylaxis, lymphadenopathy | Expected reactogenicity: injection site pain, fatigue, headache, muscle pain, joint pain and chills. Rare lymphadenopathy and hypersensitivity. | NA | Not different from placebo arm. No anaphylaxis | Injection site reactions, flu-like illness, headache, asthenia. No anaphylaxis | Expected reactogenicity: injection site pain, fatigue, headache, myalgia and fever. No anaphylaxis | NA | Not different from placebo arm. No anaphylaxis |

\*Patients included in published interim analysis MOA: Mechanism of action, PRD: Patients with rheumatic disease NA: Not available

There are currently no available data on the immunogenicity and efficacy of COVID-19 vaccination in PRD.

#### 3. Are other (non-COVID-19) recommended non-live vaccines safe, immunogenic and efficacious in PRD?

#### 4. What is the effect of various drugs used in PRD on immunogenicity of (non-COVID-19) vaccines in PRD?

To review the evidence in non-live, non-COVID-19 vaccinations in PRD, a systematic review of international best practice guidelines and recommendations from rheumatology societies on vaccinations in PRD was performed, in lieu of a systematic review of the primary literature. We searched PubMed for publications using the Medical Subject Headings (MeSH) terms (“Consensus”[MeSH] OR “Consensus Development Conference, NIH” [Publication Type] OR “Consensus Development Conference” [Publication Type] OR “Consensus Development Conferences, NIH”[MeSH] OR “Consensus Development Conferences”[MeSH]) OR (“Guidelines as Topic” [MeSH] OR “Practice Guidelines as Topic” [MeSH] OR “Guideline” [Publication Type] OR “Health Planning Guidelines” [MeSH] OR “Standard of Care” [MeSH] OR “Practice Guideline” [Publication Type] OR “Clinical Protocols” [MeSH] AND ((vaccine[MeSH Terms]) OR (vaccination[MeSH Terms])) OR (active immunization[MeSH Terms]) AND ((((autoimmune disease[MeSH Terms]) OR (rheumatology[MeSH Terms])) OR (host, immunocompromised[MeSH Terms])) OR (immunocompromised host[MeSH Terms])) OR (immunocompromised patient[MeSH Terms]). The filters English (language) and Humans were applied. This search yielded 191 citations. One member of the CWG (ML) screened through the titles and/or abstracts and excluded those that were not a practice guideline, not targeted to PRD, only addressed live vaccines, were only targeted to childhood vaccines, did not undertake a systematic literature review, were duplicates, or were outdated recommendations from the same body. Four additional citations were added from manual search. We then reviewed the remaining 21 full text articles and excluded best practice guidelines that did not undertake a consensus methodology and evidence grading or strength of recommendations. Eleven full text articles were finally included (Figure 1, Table 2).^21-31^ The definitions of PRD and immunomodulatory drugs considered in this recommendation are summarized in Table 3.

**Table 2:** Reviewed Practice Guidelines Citations, with focus on non-live vaccinations.

| Article | Vaccine type | Patient Population | Safety | Immunogenicity | Efficacy | Timing / DMARD Cessation | Post vaccination Antibody Testing | Vaccination of Household Contacts |
| --- | --- | --- | --- | --- | --- | --- | --- | --- |
| Furer V, et al. <sup>29</sup> | Non-live | PRD on IS/ DMARD/ GC | Influenza (LOE<sup>a</sup> 2b-4) and PCV (LOE 4) deemed safe | Good for Influenza (LOE 1b-4) and PPSV23 (LOE 1b-4)<br>Influenza: reduced by RTX, ABA<br>PPSV23: reduced by RTX, ABA, TOF, GOL<br>PCV13: reduced by MTX | Influenza (LOE 2a-5), PPSV23 (LOE 1b-4)<br>No data for MTX, TNFi, B cell depletion, belimumab, tocilizumab, abatacept, tofacitinib, glucocorticoids | Quiescent dx<br>Prior to IS, in particular B cell depleting therapy (6mths post RTX, 4wks before next dose of RTX)<br>No DMARD cessation | - | Yes, except for oral polio (LOE NA) |
| Seo YB, et al. <sup>22</sup> | Non-live | PRD on IS/ DMARD/ GC | Similar risk as general population (Influenza LOE<sup>b</sup> : mod; Pneumococcal LOE : low) | Similar or slightly lower than that of healthy individuals.<br>Pneumococcal: reduced by MTX, RTX, ABA | Influenza and pneumococcal | Stable dx (LOE : very low)<br>Prior to IS (LOE : very low)<br>Before ABA and ≥4 wks before RTX<br>No DMARD cessation | - | Yes |
| Guerrini G, et al. <sup>28</sup> | Influenza and pneumococcal | PRD on IS/ DMARD/ GC | Influenza and Pneumococcal deemed safe (LOE<sup>c</sup> 2) | Pneumococcal: reduced by MTX, RTX, ABA, TOF, MMF, AZA, CyC, high dose GC (LOE 2)<br>Influenza: reduced by RTX, ABA, high dose GC (LOE 2) | - | Stable dx (LOE 2)<br>Pneumococcal: before IS and ≥4 wks before RTX (LOE 2) | - | - |
| Papp KA, et al. <sup>24</sup> | Non-live | PRD on IS/ DMARD/ GC | - | - | - | 2 wks before IS (LOE<sup>d</sup> mod)<br>RTX: 5 mths post RTX and ≥4 wk prior to RTX (LOE low) | - | - |
| Holroyd CR, et al. <sup>26</sup> | Non-live | RA, PsA, axSpA | No flare of RA with Influenza | Influenza: reduced by ETN and INF, RTX, ABA<br>Pneumococcal: reduced by MTX, RTX, ABA (LOE<sup>d</sup> 1C) | - | - | - | - |
| Keeling SO, et al. <sup>25</sup> | Influenza | SLE | Trivial number of SLE flares with Influenza (LOE<sup>e</sup> mod) | - | Influenza (LOE mod) | - | - | - |
| Singh JA, et al. <sup>21</sup> | Non-live | RA on DMARD/ GC | - | Reduced by RTX and possibly MTX (LOE<sup>d</sup> very low) | Killed vaccine (LOE very low) | No DMARD cessation needed (LOE very low) |  |  |
| Bühler S, et al. <sup>30</sup> | Non-live | PRD on IS/<br>DMARD/ GC | No flare nor<br>trigger of<br>rheumatic disease,<br>(LOE<sup>d</sup> low) | Reduced by DMARD/<br>GC especially MTX,<br>RTX, ABA (LOE mod) | - | When the IS is lowest (LOE low)<br>Before ABA<br>RTX: 6 mths post RTX for<br>revaccination, 12 mths post RTX for<br>primary vaccination | 4-6 wks post<br>vaccine (LOE NA) | Yes (LOE NA) |
| Rubin LQ, et al. <sup>23</sup> | Non-live | IC | - | Influenza: reduced<br>within 6 months of RTX<br>(LOE<sup>5</sup> mod) | - | ≥2 wks before IS (LOE mod) | - | Yes (LOE high) |
| Centers for Disease<br>Control & Prevention<br><sup>31</sup> | Pneumococcal | IC | - | - | - | - |  |  |
| Heijstek MW, et al. <sup>27</sup> | Non-live | PRD on DMARD<br>/GC | No flare of<br>rheumatic disease<br>or serious adverse<br>events in<br>comparison to<br>healthy subjects | Influenza: reduced by<br>GC >10mg/day (LOE<sup>c</sup><br>3), AZA, HCQ, CYC<br>(LOE 2), RTX (LOE 2)<br>Pneumococcal: reduced<br>by MTX (LOE 2), RTX<br>(LOE 1b) | - | Before RTX (LOE 1b-2) | Influenza and<br>pneumococcal: On<br>RTX (LOE 1b-2),<br>GC ≥2mg/kg or<br>20mg/d for ≥2wks<br>(LOE 3), ±TNFi<br>(LOE 2)<br>PPSV23: On MTX<br>(LOE 2) | - |

a. Oxford Centre for Evidence-Based Medicine – levels of evidence <sup>46</sup>.
b. Level of evidence as defined: High - Very unlikely to change confidence in the estimate of effect by an additional study; Moderate - Likely to change confidence in the estimate of effect by an additional study; Low - Highly likely to change confidence in the estimate of effect by an additional study; Very low - Not sure about confidence in the estimate of effect
c. Level of evidence as defined: 1a - Meta-analysis of randomised controlled trials (RCT); 1b - Randomised controlled trials; 2 - Prospective controlled intervention study without randomization; 3 - Descriptive/analytic study (including case-control, cross-sectional, case series); 4 - Expert committee reports or opinion or clinical experience of respected authorities or both
d. GRADE level of evidence <sup>32</sup>.
e. Level of evidence as defined: High - Consistent evidence from well performed RCTs or exceptionally strong evidence from unbiased observational studies; Moderate - Evidence from RCTs with important limitations (inconsistent results, methodological flaws, indirect, or imprecise) or exceptionally strong evidence from unbiased observational studies; Low - Evidence for at least 1 critical outcome from observational studies, RCTs with serious flaws or indirect evidence; Very low - Evidence for at least 1 critical outcome from unsystematic clinical observations or very indirect evidence
Aza: azathioprine; ABA: abatacept; axSpA: axial spondyloarthritis; CYC: cyclophosphamide; dx: disease; DMARD: disease modifying anti-rheumatic drugs; GC: glucocorticoid; IS: immunosuppression; IC: immunocompromised; JAKi: inhibitors of janus kinase; PsA: psoriatic arthritis; SLE: systemic lupus erythematosus; LOE: level of evidence; mths: months; MTX: methotrexate; MMF: mycophenolate mofetil; mod: moderate; NA: non-available; PRD: people with rheumatic diseases; PCV: pneumococcal vaccination; RA: rheumatoid arthritis; RTX: rituximab; TNFi: tumor necrosis factor inhibitors; TOC: tocilizumab; wks: weeks.

**Table 3:** Definition of PRD and Immunomodulatory treatment

|  |
| --- |
| PRD include, but are not limited to, those diagnosed with:<br>1. Chronic inflammatory arthritides (e.g. rheumatoid arthritis, psoriatic arthritis, spondyloarthritides, juvenile idiopathic arthritis, adult onset stills disease)<br>2. Connective tissue diseases (e.g. systemic lupus erythematosus, immune mediated inflammatory myositis, sjögrens syndrome, systemic sclerosis)<br>3. Primary systemic vasculitides<br>4. Autoinflammatory diseases |
| Immunomodulatory drugs considered for this guidance include:<br>1. Conventional synthetic disease modifying anti-rheumatic drugs (DMARDs) (methotrexate, sulphasalazine, leflunomide, hydroxychloroquine)<br>2. Biologic DMARDs (anti-tumour necrosis factor, tocilizumab, rituximab, abatacept, secukinumab, ixekizumab, anakinra, belimumab)<br>3. Targeted synthetic DMARDs (tofacitinib, baricitinib, upadacitinib*)<br>4. Immunosuppressive drugs (cyclophosphamide, mycophenolate mofetil, azathioprine, cyclosporin A, tacrolimus)<br>5. Glucocorticoids<br>*not included in any of the searched literature on vaccines, hence recommendation is by extrapolation |

**Figure 1:**
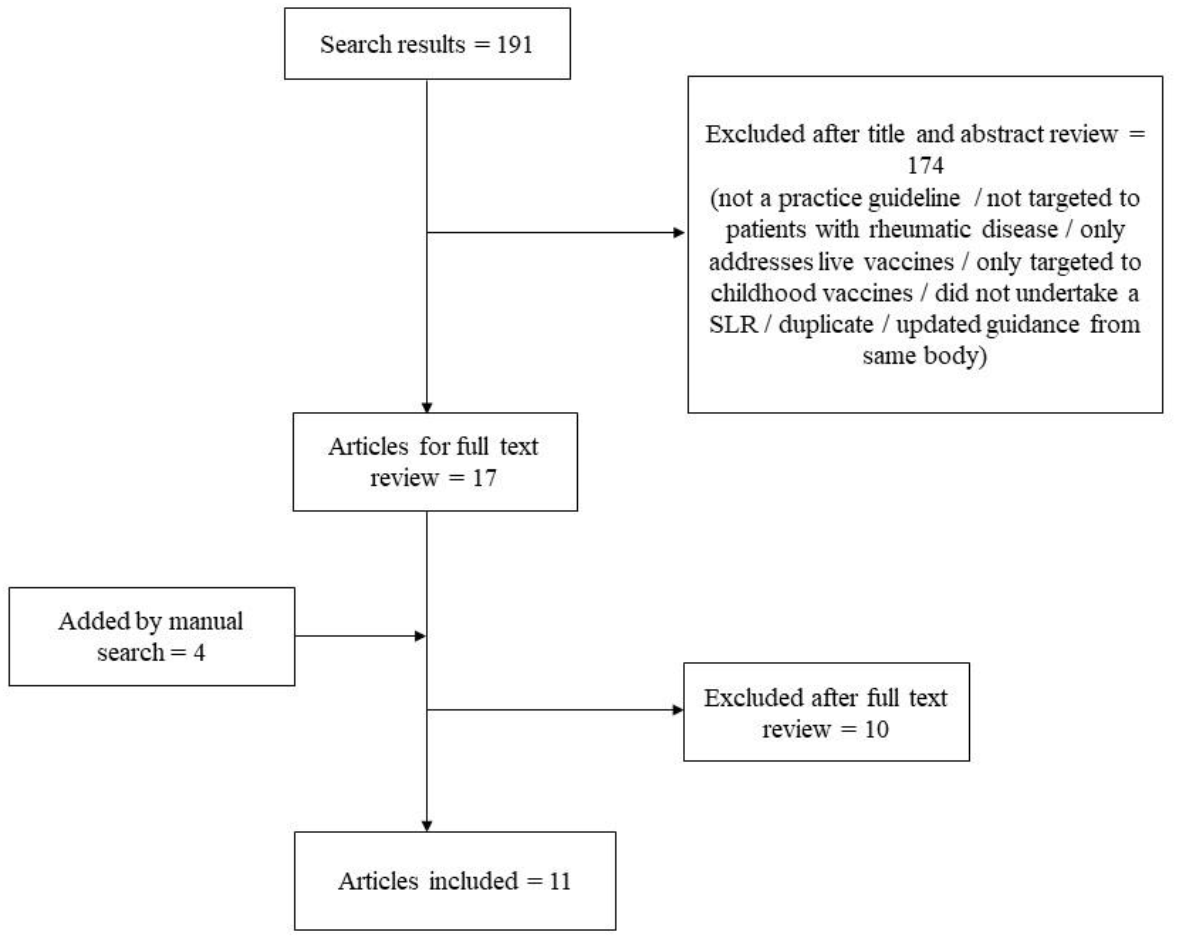
Flowchart for study selection.

### Creation of preliminary statements and rating

The CWG met to formulate and finalize preliminary statements for rating by the TFP, which was conducted on an online survey platform. The TFP were provided with summarized evidence from the reviewed trials and guidelines, a link to an online rating form and rating instructions. Based on their expertise and the provided literature, each TFP member independently rated each statement on a five-point Likert scale (1 = strongly disagree, 2 = disagree, 3 = neutral, 4 = agree, 5 = strongly agree); an agreement was defined as a score of 4 or 5. A consensus was obtained if there was ≥70% agreement. The CWG and the TFP convened via a teleconferencing platform, where the aggregated findings were presented and discussed. Definitions were clarified and statements were reworded, if needed. As there was consensus on all statements following the online voting round, no further round of voting was conducted. The Grading of Recommendations Assessment, Development and Evaluation (GRADE) methodology ^32^ was used to determine the strength of recommendations. In determining the strength of recommendations, the TFP considered the level of evidence available, as well as the balance between the potentially expected benefits and risks from COVID-19 vaccination/ omission of vaccination in PRD. Recommendations were categorized as “strong” when benefits/risks clearly outweighed the other, and “conditional” when benefits/risks were closely balanced or uncertain.

### Finalizing Consensus Statements

The final consensus statement was circulated to the TFP after the consensus meeting and was approved by all members.

## Results

The final consensus statements consist of two overarching principles and seven recommendations. They are summarized in Table 4.

**Table 4:** Final Consensus Statements

|  | Median Likert Score | % Agreement | Strength of Recommendation |
| --- | --- | --- | --- |
| **Overarching Principles** |  |  |  |
| Vaccination in people PRD should be aligned with prevailing national policy. | 4.5 | 100 | - |
| The decision for vaccination should be individualised, and should be explained to the patient, to provide a basis for shared decision-making between the healthcare provider and the patient. | 5 | 100 | - |
| **Recommendations** |  |  |  |
| 1. We *strongly* recommend that eligible patients be vaccinated against SARS-CoV2. | 5 | 100 | Strong |
| 2. We *conditionally* recommend that the COVID-19 vaccine be administered during quiescent disease, if possible. | 4.5 | 100 | Conditional |
| 3. We *conditionally* recommend that immunomodulatory drugs, other than rituximab, can be continued alongside COVID-19 vaccination. | 5 | 80 | Conditional |
| 4. We *conditionally* recommend that the COVID-19 vaccine be administered prior to commencing rituximab, if possible. For patients on rituximab, the vaccine should be administered a minimum of 6 months after the last dose, and/or 4 weeks prior to the next dose of rituximab. | 4 | 90 | Conditional |
| 5. We *conditionally* recommend that post-COVID-19 vaccination antibody titres need not be measured. | 4 | 90 | Conditional |
| 6. We *strongly* recommend that household contacts be vaccinated against SARS-CoV2. | 4.5 | 100 | Strong |
| 7. We *conditionally* recommend that any of the approved COVID-19 vaccines may be used, with no particular preference. | 4 | 70 | Conditional |

### Overarching principles

#### 1. Vaccination in PRD should be aligned with prevailing national policy

The knowledge on COVID-19 vaccination is rapidly evolving with various candidate vaccines still undergoing clinical trials. As new evidence becomes available, the landscape of vaccine availability in each country will likely differ. It is important that healthcare professionals align their recommendations to prevailing national policy, to ensure consistency of messages to patients and maintain streamlined safety workflows. Vaccine safety monitoring systems, such as the vaccine adverse event reporting system (VAERS) are in place to detect possible safety signals in the vaccinated population. Locally, the HSA reviews all reports of post-vaccination reactions, to inform national policy of vaccine eligibility, monitoring and precautions.

#### 2. The decision for vaccination should be individualised, and should be explained to the patient, to provide a basis for shared decision-making between the healthcare provider and the patient

The Rheumatologists’ decision for offering vaccinations to their patients should take into account the individual patient’s disease state, medications, as well as their risk profile and preferences. Patients should be provided with evidenced-based information to enable them to participate in a shared decision-making process. Information should include the potential risks and benefits from vaccination (or its omission), the vaccination schedule and a discussion of the various available vaccines.

### Recommendations

#### 1. We strongly recommend that eligible patients be vaccinated against SARS-CoV2

PRD are a vulnerable patient population at increased risk of acquiring COVID-19^1^ and suffering severe outcomes^3,15^. While there are little data on mRNA vaccination in PRD, there are no reports of autoimmune disease flares in the small group of PRD included in the Pfizer/BioNTech^®^ trial.^4^ There is an isolated report of a healthy individual who was diagnosed with fatal immune thrombocytopenia six days after COVID-19 vaccination with no clear evidence of the development of a new autoimmune disease. While there was temporal association, it could not be fully concluded that the vaccine was definitely the cause for the patient’s presentation.^33^ To our knowledge, there are no other published reports of autoimmune disease induction or flare after COVID-19 vaccination in the more than 300 million people vaccinated worldwide to date. COVID-19 vaccination should therefore be strongly recommended for PRD given the vulnerability of PRD along with good efficacy, immunogenicity and favourable safety profile of COVID-19 vaccination in healthy patients. This is in line with recommendations endorsed by the British Society of Rheumatology for clinically extremely vulnerable (CEV) patients^34^, which includes PRD and the recent press release from the American College of Rheumatology (ACR)^35^. The United States Centers for Disease Control and Prevention (US CDC) similarly places immunocompromised persons at an increased risk for severe COVID-19 and recommends that these patients receive vaccination as long as there are no contraindications^.36^

#### 2. We conditionally recommend that the COVID-19 vaccine be administered during quiescent disease, if possible

This recommendation is extrapolated from other vaccine recommendations in PRD, and is largely based on expert opinion, hence the conditional strength of recommendation. Vaccination studies in PRD have been largely conducted during quiescent disease state ^28,29^ and thus have limited generalizability to the PRD population with active disease, though isolated studies have shown similar vaccine immunogenicity regardless of disease state ^37^. The decision for vaccination in patients whose disease is not quiescent should be considered on an individual patient level.

#### 3. We conditionally recommend that immunomodulatory drugs, other than rituximab, can be continued alongside COVID-19 vaccination

Vaccination studies in PRD on immunomodulatory drugs (other than B cell depleting therapy) have shown sufficient protective efficacy with common non-live vaccines including influenza and pneumococcal vaccines, despite somewhat reduced immunogenicity particularly with methotrexate and abatacept^.22,26,30^

#### 4. We conditionally recommend that the COVID-19 vaccine be administered prior to commencing rituximab, if possible. For patients on rituximab, the vaccine should be administered a minimum of 6 months after the last dose, and/or 4 weeks prior to the next dose of rituximab

B cell depleting therapy with rituximab is associated with significant reduction in immunogenicity. Despite reduced humoral immune response, cellular immune response is still preserved after influenza vaccination in patients who were treated with rituximab.^38^ Satisfactory immunogenicity has been shown in rituximab treated patients when influenza and pneumococcal vaccines were administered 6 months after a previous dose ^23,29,39^ and at least 4 weeks prior to a subsequent dose ^24,28^, forming the basis of this conditional recommendation. Of note, the British Arthritis and Musculoskeletal Alliance recommends that vaccination should not be delayed in patients on or planned for rituximab, with an ideal interval of vaccination 4-8 weeks after the last dose of rituximab or 2 weeks prior to a planned dose of rituximab, if possible^.34^

#### 5. We conditionally recommend that post-COVID-19 vaccination antibody titres need not be measured

Outside of paediatric care ^27^, post-vaccination antibody titre measurement is not part of routine clinical practice and is not part of other vaccination guidelines in adult PRD. As the correlation between antibody titres post COVID-19 vaccination and clinical protection is not well established at present, we conditionally recommend that titres not be measured.

#### 6. We strongly recommend that household contacts be vaccinated against SARS-CoV-2

Vaccination of household contacts has been advocated by societies such as European Alliance of Associations for Rheumatology (EULAR) ^29^ and Infectious Diseases Society of America (IDSA) ^23^ for a variety of inactivated and live vaccines (except for the oral polio vaccination ^22,23,29^). Increasingly, epidemiologic studies have demonstrated SARS-CoV-2 transmission in close contacts due to asymptomatic and pre-symptomatic infections ^40-42^, highlighting the importance of extending vaccinations to household contacts in order to protect vulnerable patients.

#### 7. We conditionally recommend that any of the approved COVID-19 vaccines may be used, with no particular preference

The various SARS-CoV-2 vaccines in development are non-live vaccines. The anticipated risk benefit ratio should therefore be similar for vaccinations to be recommended without preference for any particular vaccine. However, long term follow-up in PRD will be needed to ascertain longer term efficacy and safety of the various vaccines.

## Discussion

The consensus recommendations for COVID-19 vaccination in PRD presented in this article were based on review of the limited currently available literature with these vaccines, supplemented by the more extensive knowledge that is available for other non-live vaccines in PRD. It is noteworthy that the absence of evidence is not evidence of absence, and practical recommendations for PRD need to be made despite the scarcity of literature in these vulnerable patients. Experts in the specialty were consulted, in order to synthesize the available literature into clinically meaningful recommendations. Available evidence on the risk of COVID-19 in PRD was weighed against the potential risks / benefits of vaccination with a new vaccine technology, borrowing from the principles of vaccination with non-live viruses in PRD and the available knowledge on mRNA drug delivery systems.

In formulating these recommendations, the TFP were cognizant of the heightened risk of COVID-19 in our patients. Therefore, recommendations were formulated to aid practicing rheumatologists in their decision making without being overly restrictive, while allowing individualized decision making for each patient. These should take into account patient’s disease status, ongoing treatment, risk profiles, preferences and local community transmission risk.

Our consensus recommendations for COVID-19 vaccinations in PRD were developed employing a systematic literature review and Delphi method. The process of recommendation development incorporated all components of the Appraisal of Guidelines for Research & Evaluation (AGREE) instrument^43^, other than patient/ allied health involvement, for practicality. The AGREE framework was developed to ensure the rigor of guideline formulation which are feasible for clinical practice. The only other consensus recommendations developed using a standardised Delphi method for COVID-19 vaccination in PRD were recently announced in a press release by the American College of Rheumatology^35^. Importantly, the broad principles for COVID-19 vaccination in PRD in our recommendations are similar to what the ACR has outlined, in spite of the vastly different pandemic situation (and therefore the balance of risk/ benefit of the vaccine) in Asia versus North America. Vaccination is strongly encouraged, may be given while on immunomodulatory therapy, preferably during quiescent disease, and without the need for testing for post-vaccination antibody titres. The ACR recommended that COVID-19 vaccination should be timed according to the dosing of certain immunomodulatory treatments (rituximab, intravenous abatacept and intravenous cyclophosphamide) and that treatment with methotrexate, Janus Kinase inhibitors and abatacept should be temporarily interrupted prior to or after COVID-19 vaccination. However, as discussed, while there may be reduced vaccine immunogenicity in patients on these medications, sufficient protective efficacy has been demonstrated^22,26,30^, thus forming the basis of our recommendation to vaccinate without treatment interruption or consideration for timing of doses.

As of the latest WHO update on March 5^th^, 2021, 79 candidate vaccines are in clinical development, with a further 182 in pre-clinical development.^44^ Since the roll out of vaccination campaigns in various regions in mid-December 2020 up to March 9^th^, 2021, more than 312 million vaccine doses have been administered worldwide^45^ and our collective experience with the new vaccines continue to evolve. It is important that governing institutions and healthcare providers continue to keep abreast of the latest evidence, so that recommendations can be reviewed and/or revised as new knowledge emerges. Particularly, data on safety and efficacy of vaccination in PRD are urgently needed to update recommendations in this vulnerable population.

## Data Availability

This is a consensus statement on the recommendations for COVID19 vaccination in people with rheumatic disease. There are no raw data attached to this manuscript.

## Notes

### Competing Interest Statement

The authors have declared no competing interest.

### Funding Statement

No funding was received for this work.

### Author Declarations

NA. Consensus recommendations by literature review and expert panel. No IRB needed.

### Summary of Updates

The manuscript has been updated to keep up with the rapidly evolving field of vaccine development. Approved vaccines and global statistics have been updated to be as current as possible.

